# Minocycline Transforms Acute Stroke Care: A 2,428-Patient Meta-analysis of a Low-Cost Neuroprotective Strategy

**DOI:** 10.64898/2026.06.24.26356506

**Authors:** Mario Saul Lira Castañeda, Natanael Duarte, Yovanny Solorio, María Fernanda Gutiérrez Aguilera, Amir Hjeala-Varas, Maria Alejandra Rossell Ulloa, Shashvat Desai, Neel S. Singhal

**Affiliations:** Universidad Juárez del Estado de Durango, Facultad de Medicina y Nutrición, Durango, México; Holy Name Medical Center, Department of Medicine, Teaneck, New Jersey, USA; Community Regional Medical Center, Department of Neurosurgery, Fresno, California, USA; Universidad 0de Guanajuato, Departamento de Medicina y Nutrición, Guanajuato, México; Universidad Católica Boliviana San Pablo, Santa Cruz, Bolivia; Universidad Privada del Valle, Cochabamba, Bolivia; HonorHealth, Department of Neurology, Scottsdale, Arizona; University of California San Francisco, Department of Neurology, California, USA

**Keywords:** acute ischemic stroke, minocycline, neuroprotection, meta-analysis

## Abstract

**Background:** Stroke remains a leading cause of death and long-term disability worldwide, and a substantial proportion of patients experience incomplete recovery despite modern reperfusion strategies. Minocycline is an inexpensive, widely available agent with anti-inflammatory, anti-apoptotic and matrix metalloproteinase-modulating properties that make it an attractive neuroprotective adjunct in acute ischemic stroke or intracerebral hemorrhage. However, prior clinical studies have been individually underpowered or methodologically heterogeneous, underscoring the need for an updated quantitative synthesis focused on clinically meaningful outcomes.

**Methods:** This systematic review and meta-analysis was conducted to evaluate the efficacy and safety of minocycline in adults with AIS and ICH. The search was completed on February 24, 2026, and the protocol was prospectively registered in PROSPERO (CRD420261354283). A total of 1,633 records were screened. A total of 11 studies comprising 2428 patients were included in the review dataset. Outcomes were analyzed including 90-day disability, neurological recovery, recurrent stroke, and composite vascular events (cardiovascular event, non-fatal stroke, and non-fatal MI).

**Results:** Minocycline was associated with better 90-day neurological recovery, with a greater reduction in NIHSS score at 90 days (mean difference [MD] −2.17, 95% CI −2.68 to −1.65, moderate certainty) and lower functional disability at 90 days measured by mean modified Rankin Scale (mRS) score (MD −0.25, 95% CI −0.38 to −0.13, moderate certainty). Categoric functional outcomes also favored minocycline, including mRS 0-1 at 90 days (odds ratio [OR] 1.21, 95% CI 1.02 to 1.45, high certainty), while the effect for mRS 0-2 at 90 days was borderline (OR 1.21, 95% CI 1.00 to 1.47, moderate certainty). No significant difference was observed for stroke recurrence at 90 days (OR 1.13, 95% CI 0.78 to 1.64). Composite vascular events at 90 days also favored minocycline (OR 1.21, 95% CI 1.02 to 1.45).

**Conclusions:** Minocycline appears to be a promising, scalable, and low-cost adjunctive therapy for acute ischemic stroke and intracerebral hemorrhage with evidence of improved 90-day functional and neurological outcomes. Given its global availability, favorable biological rationale, and consistent direction of effect across key recovery outcomes, these findings support prioritization of minocycline for confirmatory trials and highlight its relevance in stroke and neurocritical care clinical practice.

## INTRODUCTION

Stroke is the second leading cause of death and the third leading combined cause of death and disability worldwide, accounting for approximately 12.2 million new cases and 6.6 million deaths annually (1). Global Burden of Disease Study 2024 analysis reported that acute ischemic stroke (AIS) alone was responsible for over 7 million deaths and more than 160 million disability-adjusted life-years in 2021, with incidence disproportionately concentrated in low- and middle-income countries (1,2). Despite major advances in reperfusion therapy, including intravenous thrombolysis and mechanical thrombectomy, these interventions are time-dependent and available to only a minority of patients globally; a substantial proportion of survivors are left with permanent disabilities (3). Adjunctive neuroprotective strategies capable of extending the therapeutic window and attenuating secondary injury, therefore, remain an important and unmet priority in contemporary acute stroke care (3,4). In addition, targeting secondary neurological injury mediated by cell death programs, neuroinflammatory pathway, and matrix metalloproteinase (MMP)-mediated extracellular matrix degradation (4) remains a critical clinical therapeutic gap to prevent infarct expansion, hemorrhagic transformation, and promote long-term neurological recovery (5).

Minocycline, a second-generation tetracycline with favorable BBB penetration and an established clinical safety record, has emerged as a compelling repurposed candidate for AIS owing to its pleiotropic neuroprotective properties (5,6). Preclinical models have shown that minocycline suppresses microglial activation, attenuates the post-ischemic neuroinflammatory response, and reduces neuronal apoptosis, resulting in smaller infarct volumes and improved functional recovery (5,6). At the vascular level, minocycline modulates MMP-mediated neurovascular injury, including inhibition of MMP-9 activity, and disrupts smooth muscle cell migration through the ERK1/2, PI3K, and MMP signaling axes, mechanisms that are directly relevant to BBB integrity and hemorrhagic transformation after stroke (7,8,9).

Early phase II trials demonstrated that oral minocycline administered within 24 hours of AIS onset was associated with improved neurological outcomes at 90 days (10). A 2018 systematic review and meta-analysis of randomized trials including 426 patients found that minocycline was associated with greater 90-day functional independence and lower NIHSS scores in patients with AIS and intracerebral hemorrhage (ICH) but concluded that the evidence base remained insufficient for definitive recommendations (11). The recently completed EMPHASIS trial, a multicenter double-blind randomized controlled trial enrolling 1,724 patients, subsequently demonstrated that minocycline initiated within 72 hours of AIS onset significantly improved the probability of excellent functional outcome at 90 days without an excess safety signal (12). Nonetheless, the broader clinical evidence base remains fragmented by heterogeneity in treatment timing, dosing, stroke severity, background reperfusion therapy, and outcome definitions, which limits the interpretability of individual studies (11,12).

Given minocycline’s low cost and global availability, position it as a potentially scalable therapy relevant even in resource-constrained settings where advanced reperfusion infrastructure remains inaccessible (3,11). In the present systematic review and meta-analysis, we synthesized available comparative evidence from 11 studies comprising 2,428 patients to evaluate the efficacy and safety of minocycline in adults with stroke, with a primary focus on clinically meaningful 90-day neurological and functional outcomes.

## METHODS

### Study design and protocol registration

This study is reported in accordance with the Preferred Reporting Items for Systematic Reviews and Meta-Analyses (PRISMA) guidelines (13). This review was prospectively registered to PROSPERO, an international prospective register of systematic reviews (ID: CRD420261354283).

### Literature search strategy

A systematic literature search was performed on February 24, 2026, in the databases PubMed, EMBASE, Web of Science, and Cochrane, using free and controlled vocabulary for the terms “stroke” and “cerebrovascular disease” and “cerebral ischemia” and “intracerebral hemorrhage” and “hemorrhagic stroke” and “minocycline.” Table S1 in the supplementary material presents the complete search strategy. No filters were applied. We also searched the reference lists of the included papers to identify additional studies reporting on the effect of minocycline on stroke, using a snowball technique.

### Eligibility criteria

This review required articles to meet the following criteria:

Human subjects aged 18 years or older were included.

1. Patients had a confirmed diagnosis of AIS or ICH by brain CT or MRI within 72 h of symptom onset
2. Studies evaluated the use of minocycline as adjuvant treatment for stroke patients.
3. Any route of minocycline administration (intravenous, intra-arterial, intrathecal, or others) was considered.
4. Studies included a comparator group receiving the best medical treatment or usual care.
5. Randomized controlled trials and cohort studies (prospective or retrospective) were included.
6. Studies reported at least one relevant clinical outcome (e.g., NIHSS, stroke recurrence, functional outcomes such as modified Rankin Scale, or mortality).

Articles meeting the following criteria were excluded from our review:

1. Patients younger than 18 years of age.
2. Studies including AIT or unclear diagnosis of AIS or ICH
3. Studies without a comparator group.
4. Case-control studies, case reports, case series, or single-arm interventional studies.
5. Studies with overlapping patient populations (in such cases, the most complete or recent dataset was selected).
6. Articles without sufficient data on outcomes of interest.

### Data management

We used a Systematic Review Accelerator (SRA) for deduplication of the search strategy results (14). The remaining studies were imported into the web-based screening tool “Rayyan” for the screening process.

### Study selection

All article abstracts were screened in duplicate (ND/ML) in a blinded fashion via an artificial intelligence electronic tool for systematic literature reviews (https://www.rayyan.ai). Those found not complying with the inclusion criteria were removed, and any controversies were yielded by a third reviewer (YS) for consensus during an online meeting in which abstracts were reviewed. All remaining papers were screened again as a full article by at least two authors, and conflicts were settled as previously reported. A PRISMA flow diagram was constructed online with the use of the interactive R-based tool (15).

### Quality and Certainty Assessment

Risk of bias for included RCTs was assessed independently by two reviewers (ND/ML) using the Cochrane Risk of Bias tool version 2 (RoB 2) (16), evaluating five domains: randomization process, deviations from intended interventions, missing outcome data, measurement of the outcome, and selection of the reported result. Each domain was rated as “low risk,” “some concerns,” or “high risk,” with an overall risk-of-bias judgment assigned accordingly.

For non-randomized controlled studies (cohort studies), we used the Risk of Bias Due to Confounding and Bias in Selection of Participants Into the Study (ROBINS-I for non-RCTs) (17), which assesses bias across seven domains: confounding, participant selection, classification of interventions, deviations from intended interventions, missing data, measurement of outcomes, and selection of the reported result. Each domain was rated as “low risk,” “some concerns,” or “high risk,” with an overall risk-of-bias judgment assigned accordingly.

The certainty of evidence for each outcome was assessed (ML) using the Grading of Recommendations Assessment, Development, and Evaluation (GRADE) framework (18). Evidence was classified into four levels: high (we are very confident that the true effect lies close to the estimate of effect), moderate (we are moderately confident in the effect estimate; the true effect is likely to be close to the estimate, but there is a possibility it is substantially different), low (our confidence in the effect estimate is limited; the true effect may be substantially different from the estimate), and very low (we have very little confidence in the effect estimate; the true effect is likely to be substantially different from the estimate). In line with GRADE guidance, we described our results using standardized certainty language, such that, for example, minocycline “leads to improved functional status” (high certainty), “probably leads to improved functional status” (moderate certainty), “may lead to improved functional status” (low certainty), or “the effect of minocycline on improvement of functional status is very uncertain” (very low certainty), and this terminology was applied consistently throughout the manuscript.

### Data extraction

Four reviewers independently and in duplicate extracted data using a standardized, piloted form. They captured study-level variables (first author, year, country, design, number of centers, and follow-up), treatment details (sample size per arm, minocycline dose, route, timing from stroke onset, duration, concomitant reperfusion or antithrombotic therapies, and control-group regimen), and patient characteristics (age, sex, vascular risk factors, prior stroke or TIA, premorbid mRS, and stroke subtype with vascular territory or hematoma features when applicable). We additionally recorded baseline stroke severity (NIHSS and Glasgow Coma Scale when relevant), key imaging features (anterior vs. posterior circulation and thrombolysis or thrombectomy use), and the treatment window. Outcomes were extracted following a predefined hierarchy focused on 90-day disability (mRS continuous and dichotomized at 0–1, 0–2, and 0–3), neurological recovery (change in NIHSS at multiple time points), early neurological deterioration, recurrent stroke at 90 days, composite vascular events (stroke, myocardial infarction, vascular death). The safety endpoints include symptomatic intracranial hemorrhage, any bleeding and its severity, vascular and all-cause mortality, and adverse or serious adverse events potentially related to minocycline.

### Data synthesis and statistical analysis

We performed quantitative analyses in R (version 4.1.2) using the “meta” package for the quantitative synthesis of our results (19). For continuous endpoints (NIHSS and mean mRS at 90 days), we combined mean differences with 95% confidence intervals using a random-effects DerSimonian and Laird model (20) because we expected clinical and methodological differences across trials, and we showed the results in forest plots. For dichotomous functional outcomes (mRS 0–1 and mRS 0–2 at 90 days), composite vascular events at 90 days, and stroke recurrence, we calculated odds ratios with 95% confidence intervals using fixed or random effects models as appropriate (21), based on the degree of between-study heterogeneity. Heterogeneity was quantified with the I^2^ statistic and Cochran’s Q test and explored through leave-one-out sensitivity analyses to assess the influence of individual studies such as EMPHASIS and Lampl et al. on pooled estimates (22). When studies reported continuous outcomes as medians with ranges or interquartile ranges, we approximated means and standard deviations using established methods (23), assuming approximately normal distributions.

## RESULTS

The selection process is illustrated in Figure 1 (PRISMA chart flow)

**Figure 1.**
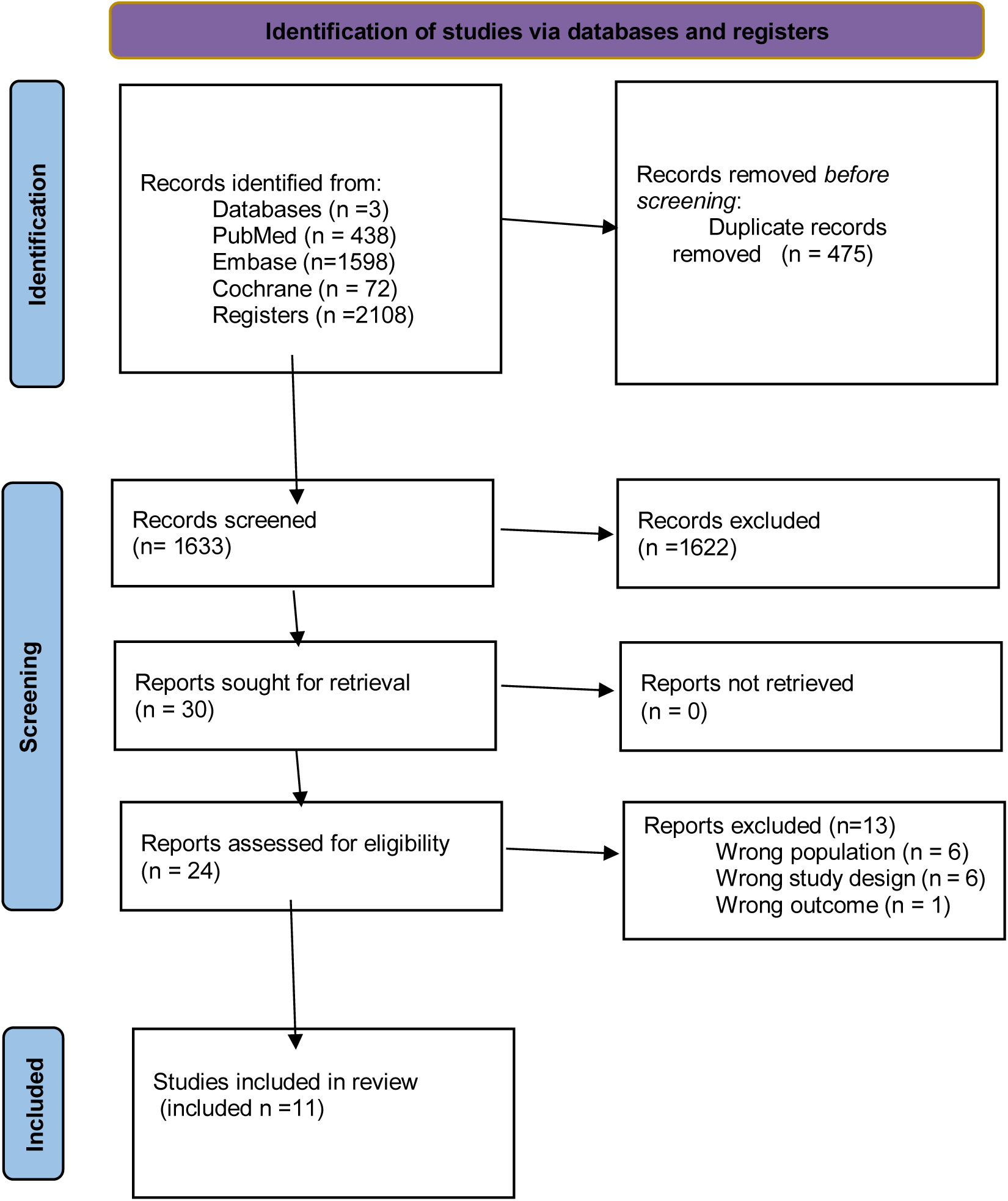
PRISMA Flow Chart

### Study Design and Stroke Type

All included studies were randomized controlled trials or prospective comparative cohorts with a control arm receiving placebo or standard care. Eight trials were double-blind or evaluator-blinded (12, 28–34), and two were single-blinded prospective studies (35,36); one was a non-randomized dose-escalation trial (37). Seven studies enrolled exclusively patients with AIS, three focused on ICH (28,32,33), and one enrolled a mixed AIS/ICH population (34). Sample sizes ranged from 16 patients (32) to 1,724 patients (12), with the EMPHASIS trial (12) alone contributing 71% of the total pooled population (Table 1)

**Table 1.**
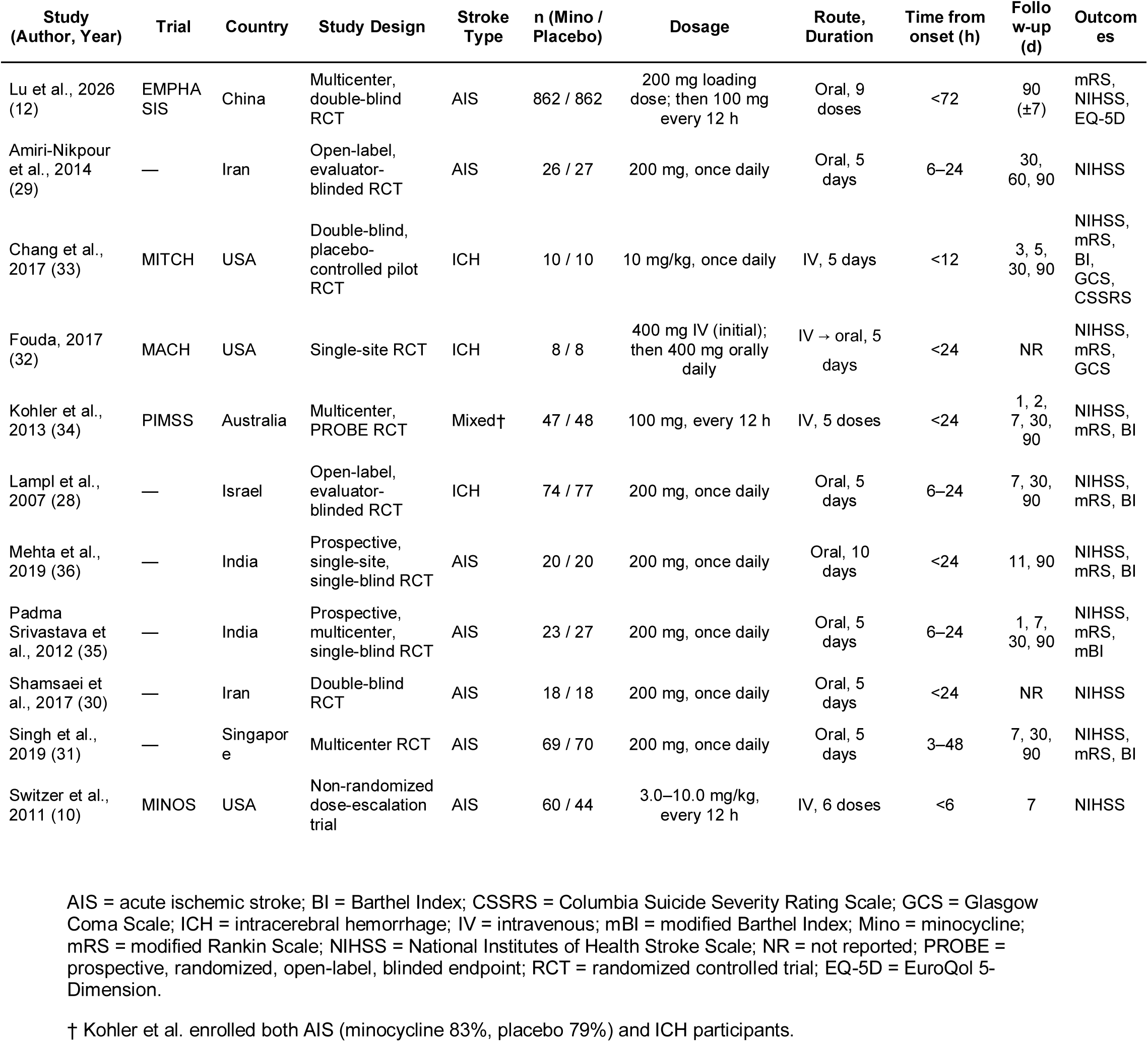
Characteristics of included studies.

### Patient demographics

The study populations were broadly similar in baseline demographics across trials. Mean age ranged from 52.7 years (35) to 69 years (30), and male sex predominated across all cohorts (range 37–75%). Hypertension was the most prevalent vascular risk factor, reported in 50–100% of patients across studies. Diabetes mellitus was present in 17–52% and dyslipidemia or hypercholesterolemia was reported in 39–56%. Baseline stroke severity, measured by the National Institutes of Health Stroke Scale (NIHSS), spanned a wide spectrum: from a median of 5 points (IQR 4–7) in the predominantly mild-to-moderate EMPHASIS cohort (12) to a mean of 13.4 (SD 4.5) in the Mehta 2019 trial, which enrolled patients with more severe strokes. Baseline characteristics were well-balanced between minocycline and control arms within each individual study (Table 2)

**Table 2.**
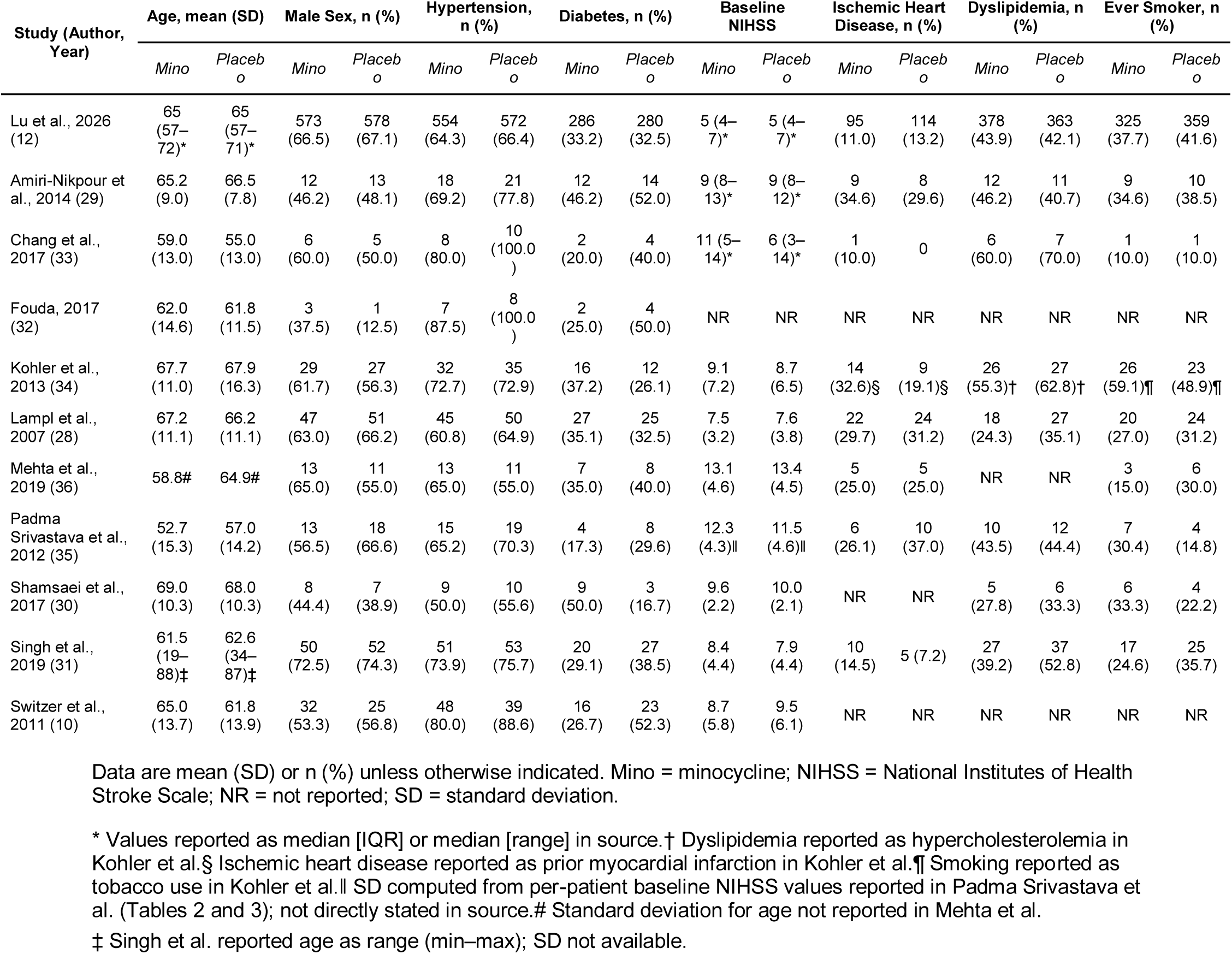
Baseline patient characteristics.

**Table 3.**
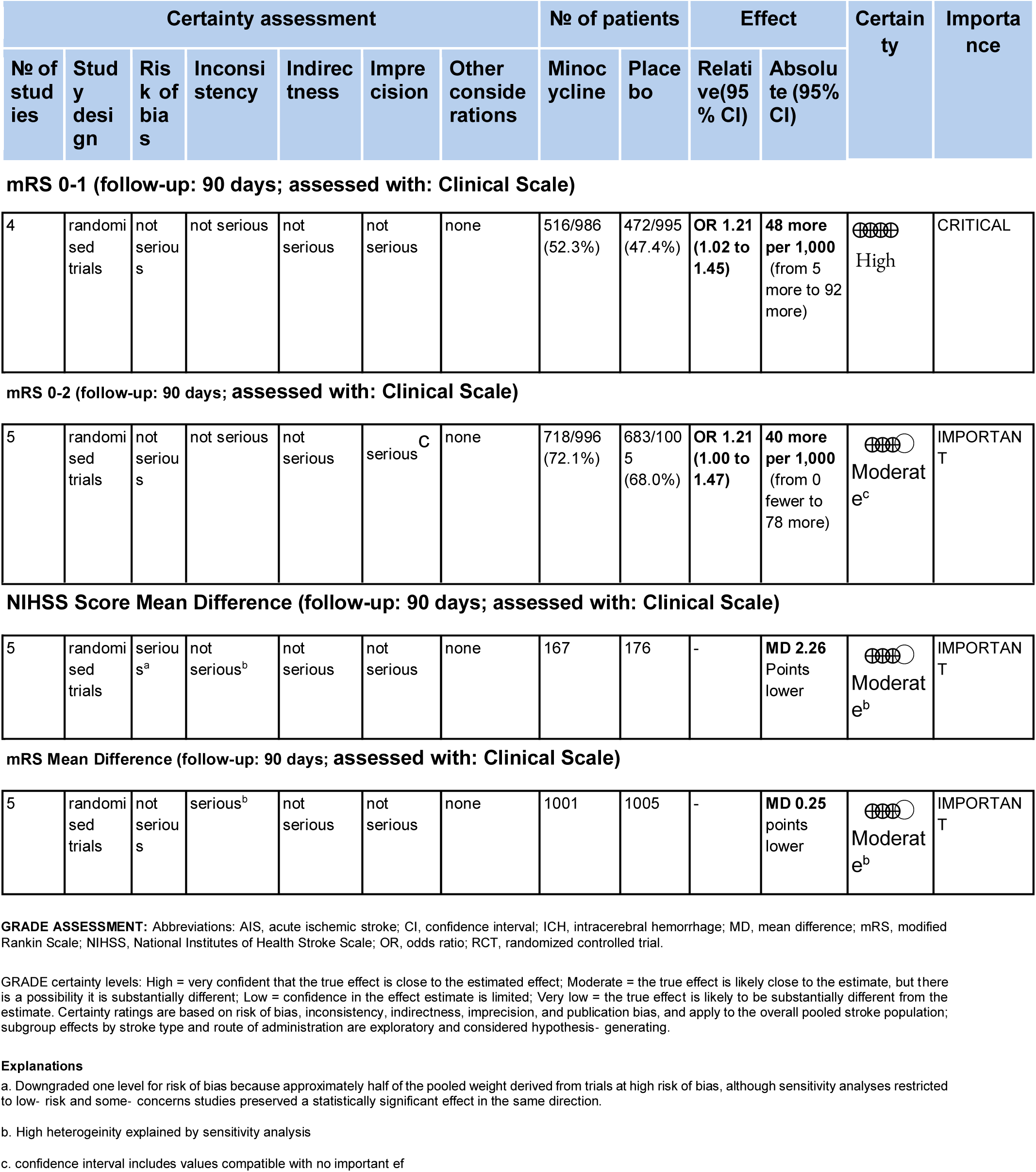
Evaluation of the quality of evidence according to Grading of Recommendations Assessment, Development and Evaluation (GRADE) working group for the primary and secondary outcomes.

| Certainty assessment |  |  |  |  |  |  | № of patients |  | Effect |  | Certainty | Importance |
| --- | --- | --- | --- | --- | --- | --- | --- | --- | --- | --- | --- | --- |
| № of studies | Study design | Risk of bias | Inconsistency | Indirectness | Imprecision | Other considerations | Minocycline | Placebo | Relative (95% CI) | Absolute (95% CI) |  |  |
| **mRS 0-1 (follow-up: 90 days; assessed with: Clinical Scale)** |  |  |  |  |  |  |  |  |  |  |  |  |
| 4 | randomised trials | not serious | not serious | not serious | not serious | none | 516/986 (52.3%) | 472/995 (47.4%) | **OR 1.21 (1.02 to 1.45)** | **48 more per 1,000** (from 5 more to 92 more) | High | CRITICAL |
| **mRS 0-2 (follow-up: 90 days; assessed with: Clinical Scale)** |  |  |  |  |  |  |  |  |  |  |  |  |
| 5 | randomised trials | not serious | not serious | not serious | serious<sup>c</sup> | none | 718/996 (72.1%) | 683/1005 (68.0%) | **OR 1.21 (1.00 to 1.47)** | **40 more per 1,000** (from 0 fewer to 78 more) | Moderate<sup>c</sup> | IMPORTANT |
| **NIHSS Score Mean Difference (follow-up: 90 days; assessed with: Clinical Scale)** |  |  |  |  |  |  |  |  |  |  |  |  |
| 5 | randomised trials | serious<sup>a</sup> | not serious<sup>b</sup> | not serious | not serious | none | 167 | 176 | - | **MD 2.26** Points lower | Moderate<sup>b</sup> | IMPORTANT |
| **mRS Mean Difference (follow-up: 90 days; assessed with: Clinical Scale)** |  |  |  |  |  |  |  |  |  |  |  |  |
| 5 | randomised trials | not serious | serious<sup>b</sup> | not serious | not serious | none | 1001 | 1005 | - | **MD 0.25** points lower | Moderate<sup>b</sup> | IMPORTANT |

**GRADE ASSESSMENT:** Abbreviations: AIS, acute ischemic stroke; CI, confidence interval; ICH, intracerebral hemorrhage; MD, mean difference; mRS, modified Rankin Scale; NIHSS, National Institutes of Health Stroke Scale; OR, odds ratio; RCT, randomized controlled trial.
GRADE certainty levels: High = very confident that the true effect is close to the estimated effect; Moderate = the true effect is likely close to the estimate, but there is a possibility it is substantially different; Low = confidence in the effect estimate is limited; Very low = the true effect is likely to be substantially different from the estimate. Certainty ratings are based on risk of bias, inconsistency, indirectness, imprecision, and publication bias, and apply to the overall pooled stroke population; subgroup effects by stroke type and route of administration are exploratory and considered hypothesis-generating.

### Risk of bias assessment

Overall, Risk of Bias 2 for randomized controlled trials showed predominantly low risk or “some concerns” across domains, with most studies judged as having appropriate randomization, minimal missing outcome data, and adequate outcome measurement, while a subset had unclear or high risk related to blinding procedures and selective reporting. ROBINS-I evaluations of non-randomized comparative studies identified moderate to serious overall risk of bias, mainly due to potential confounding, selection of participants, and deviations from intended interventions, whereas classification of interventions and outcome assessment were generally rated at lower risk. These domain-level judgments are summarized in Figure 2 and were considered when interpreting the pooled estimates in the quantitative synthesis

**Figure 2.**
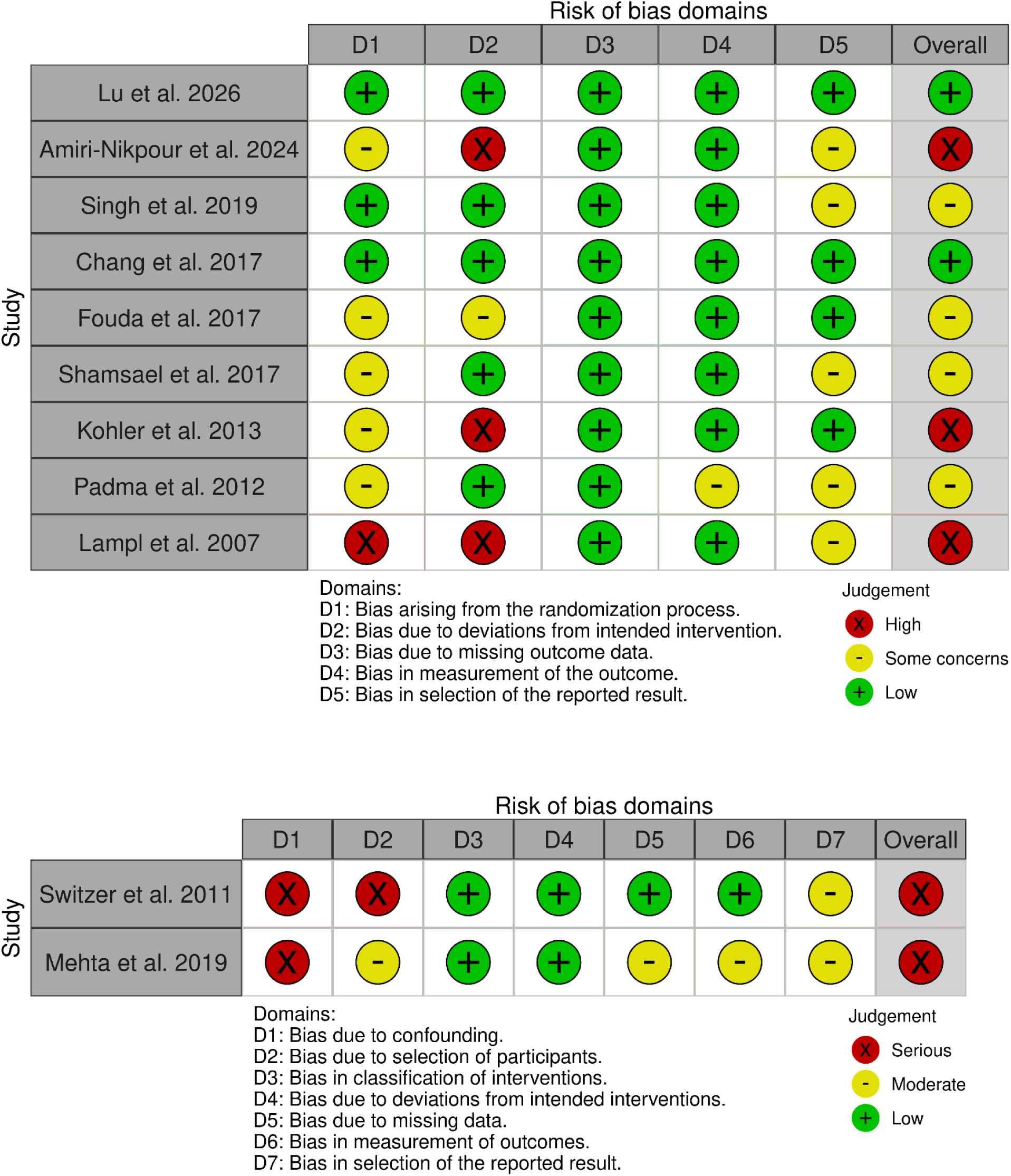
Risk of bias assessment for randomized controlled trials (RoB 2) and non-randomized studies of interventions (ROBINS-I), showing domain-specific and overall judgments for each included study.

### Minocycline Dosing and Administration

The most common regimen was oral minocycline 200 mg once daily for 5 days, used in six trials (28–31,35,36), and Mehta (36), in which treatment was extended to 200 mg daily for 10 days. The EMPHASIS trial (12) used an oral loading-dose strategy of 200 mg once, followed by 100 mg every 12 hours for 4 days. Intravenous regimens included MINOS (10) (3.0–10.0 mg/kg every 12 hours, 6 total doses), MITCH (33) (10 mg/kg/day, maximum 700 mg over 2-hour infusions for 5 days), and PIMSS (34) (100 mg IV every 12 hours for 5 doses). MACH (32) combined both routes with an initial 400 mg IV dose, followed by 400 mg orally once daily for 5 days. Across studies, treatment was started within 6–72 hours of symptom onset: ≤6 h in MINOS (10), ≤12 h in MITCH (33), 6–24 h in other studies (28,29,35), and up to 48 h (31), and primary outcomes were assessed at 90 days in all trials except MINOS (10), which reported 7-day outcomes.

### Background Therapy and Reperfusion

Use of reperfusion therapy varied substantially across studies, reflecting differences in timing and setting. In EMPHASIS (12), 45% of participants received reperfusion therapy (IV thrombolysis or mechanical thrombectomy). The PIMSS trial (34) reported IV rt-PA in approximately 15% of the patients. By contrast, Singh (31) excluded patients who received IV thrombolysis, and reperfusion therapy was not administered or reported in the remaining studies. All patients received standard stroke care in their respective settings, including antiplatelets or anticoagulation as clinically indicated. The control group received a matching placebo in double-blind trials, aspirin-based standard care in open-label comparator trials, or vitamin B complex (35).

### PRIMARY OUTCOMES

#### Neurological Recovery: NIHSS Score at 90 Days

The primary efficacy outcome was neurological recovery as measured by the change in NIHSS score at 90 days. Data were available from seven studies (n=487 patients). Minocycline probably leads to a significant reduction in NIHSS score compared with placebo or standard care (mean difference [MD] −2.17, 95% CI −2.68 to −1.65; Z=8.24, P < 0.00001; moderate certainty). Substantial heterogeneity was observed (I^2^=89%, Chi^2^=54.16, df=6, P<0.00001), driven predominantly by the Lampl et al. trial (28), which contributed 29.3% of the pooled weight and showed the largest treatment effect (MD −4.90, 95% CI −5.85 to −3.95). (Figure 3)

**Figure 3.**
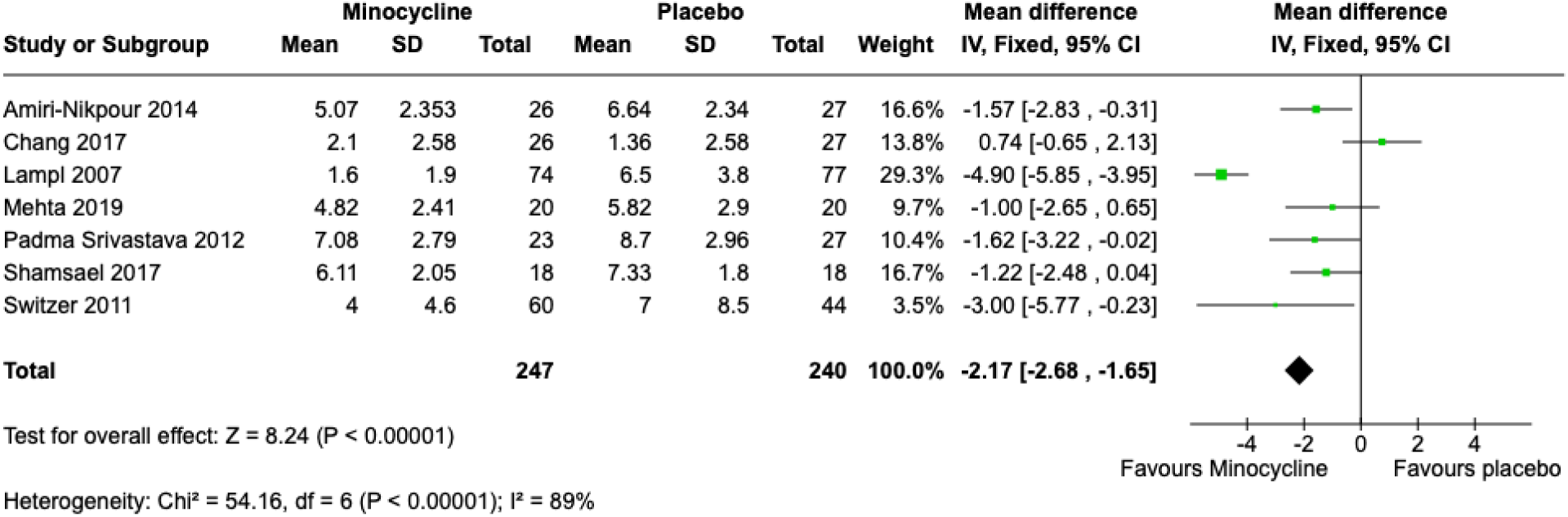
Forest plot of the mean difference in NIHSS score at 90 days, comparing minocycline with placebo in patients with acute ischemic stroke. NIHSS: National Institutes of Healt Stroke Scale

Upon exclusion of this study, the direction of effect remained consistent and statistically significant (MD −1.04, 95% CI −1.65 to −0.42; Z=3.32, P<0.00001) with attenuation of heterogeneity (I^2^=47%, Chi^2^=9.49, P=0.003). (Figure S1)

#### Functional Disability: Mean mRS Score at 90 Days

The pooled mean mRS score at 90 days was significantly lower in the minocycline group compared with placebo across five studies (n=2,006 patients). Minocycline probably leads to a significant reduction in disability (MD −0.25, 95% CI −0.38 to −0.13; Z=3.99, P < 0.0001; moderate certainty). High heterogeneity was present (I^2^=86%, Chi^2^=29.26, df=4, P<0.00001). The forest plot for the mean mRS at 90 days is shown in Figure 4.

**Figure 4.**
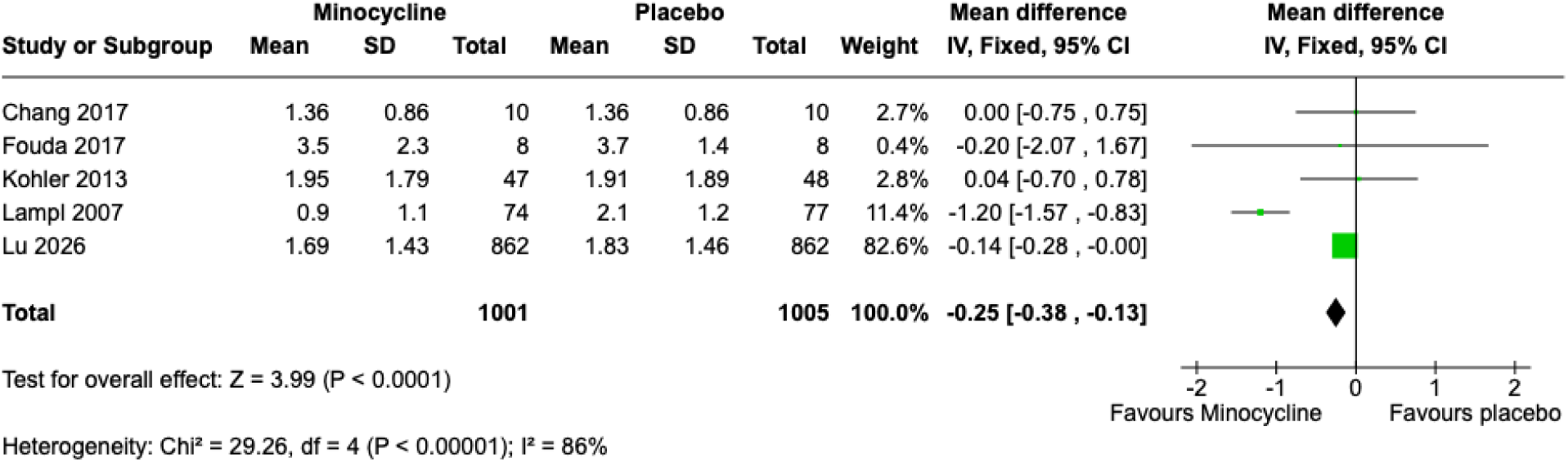
Forest plot of mean modified Rankin Scale scores at 90 days in patients treated with minocycline versus control after ischemic stroke.

In the leave-one-out sensitivity analysis excluding the EMPHASIS trial (12), the pooled effect on mean mRS at 90 days remained statistically significant and of larger magnitude (four studies, n=282; MD −0.79, 95% CI −1.08 to −0.49; Z=5.19, P<0.00001), although between-study heterogeneity increased (I^2^=79%). (Figure S2)

### SECONDARY OUTCOMES

#### Functional Independence: mRS 0–2 at 90 Days

Functional independence at 90 days was evaluable in five studies (n=2,001 patients). The pooled analysis showed a borderline significant association in favor of minocycline (odds ratio [OR] 1.21, 95% CI 1.00 to 1.47; Z=1.94, P=0.05, moderate certainty). Moderate heterogeneity was observed across studies (I^2^=52%, Chi^2^=8.31, df=4, P=0.08). In a leave-one-out sensitivity analysis excluding the EMPHASIS trial (12), the pooled effect was attenuated to the null (OR 1.03, 95% CI 0.62 to 1.72; Z=0.12, P=0.91), with persistent moderate heterogeneity (I^2^=62%, Chi^2^=7.87, df=3, P=0.05), indicating a high influence driven by EMPHASIS (12) single effect. The forest plots for the primary and sensitivity analyses are shown in Figure 5 and Figure S3, respectively.

**Figure 5.**
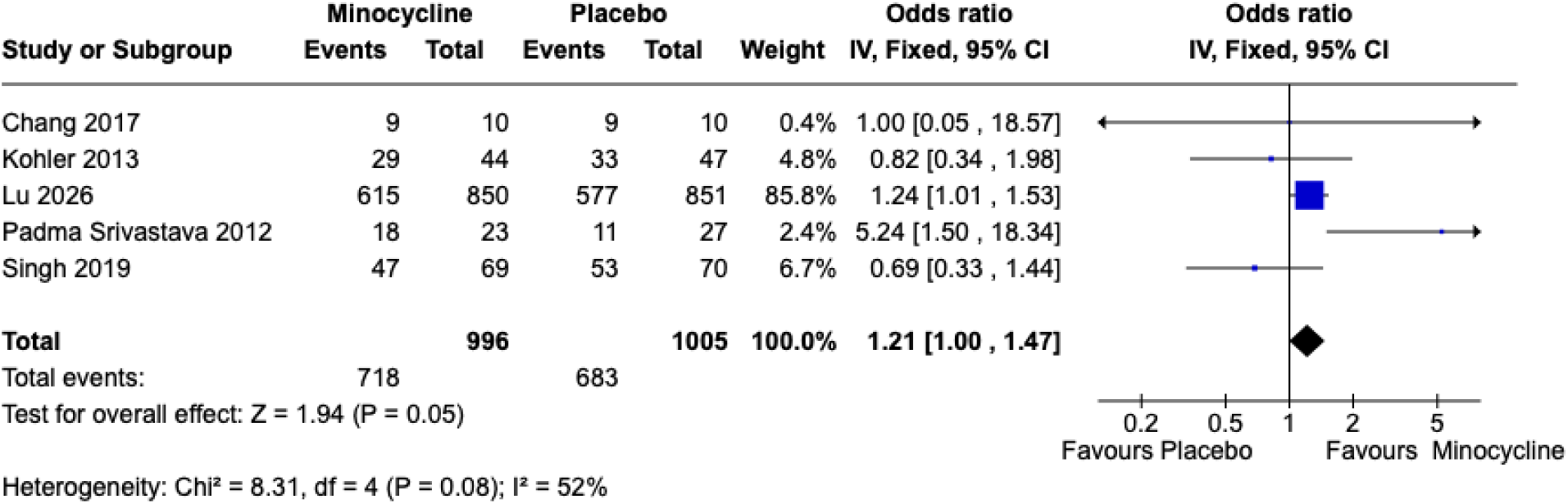
Forest plot showing the effect of minocycline versus placebo on achieving functional independence (mRS 0–2) at 90 days after ischemic stroke. mRS: modified Rankin Scale

#### Excellent Functional Outcome: mRS 0–1 at 90 Days

Functional outcome mRS 0–1 at 90 days was evaluable in four studies (n=1,981 patients). Minocycline significantly leads to a greater functional outcome compared with control (odds ratio [OR] 1.21, 95% CI 1.02 to 1.45; Z=2.14, P=0.03; high certainty). Heterogeneity was negligible (I^2^=0%, Tau^2^=0.00, Chi^2^=2.42, df=3, P=0.49). In a leave-one-out sensitivity analysis excluding the EMPHASIS trial (12), the pooled effect was attenuated and no longer statistically significant (OR 1.10, 95% CI 0.65 to 1.87; Z=0.37, P=0.71), with low residual heterogeneity (I^2^=10%, Tau^2^=0.02, Chi^2^=2.23, df=2, P=0.33). The forest plots for the main and sensitivity analyses are shown in Figure 6 and Figure S4, respectively.

**Figure 6.**
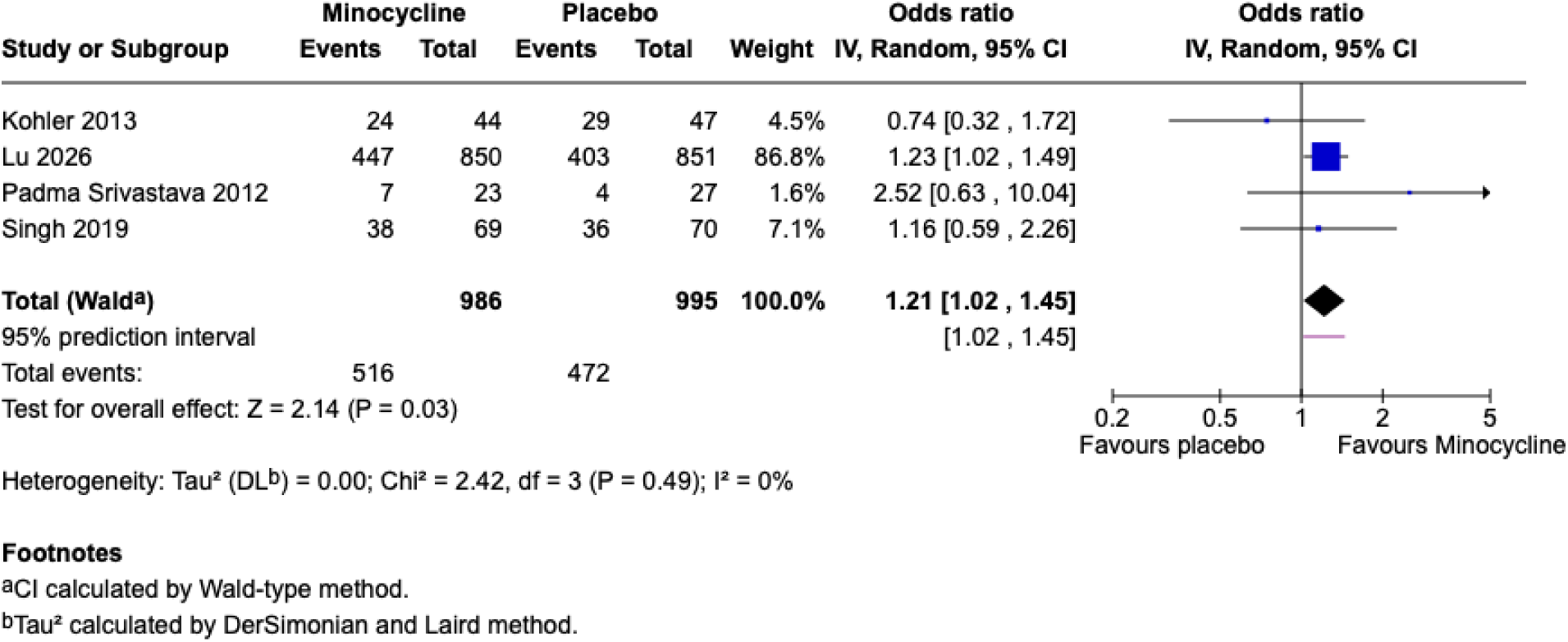
Forest plot showing the proportion of patients with an excellent functional outcome (mRS 0–1) at 90 days for minocycline versus placebo. mRS: modified Rankin Scale

#### Composite Vascular Events at 90 Days

Composite vascular events at 90 days, defined as any stroke, myocardial infarction, or vascular death, were reported in four studies (n=1,981 patients). Minocycline-treated patients probably did not significantly prevent composite vascular events compared with controls (OR 1.13, 95% CI 0.78 to 1.64; moderate certainty), with low between-study heterogeneity (I^2^=0%, Chi^2^=1.56, df=2, P=0.46). The forest plot for this outcome is presented in Figure 7.

**Figure 7.**
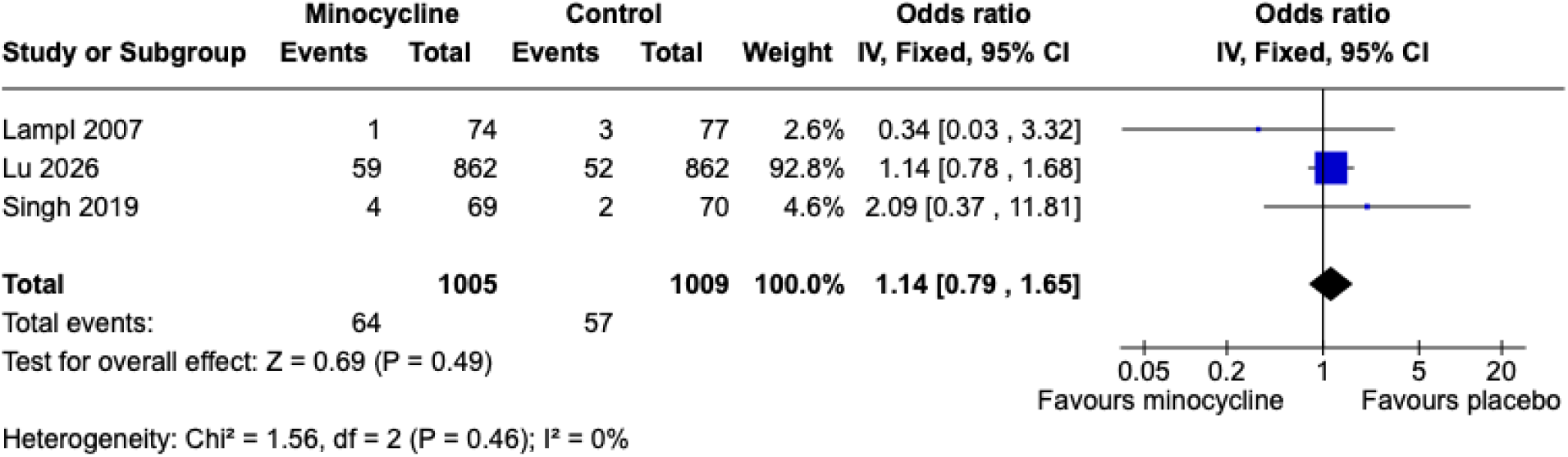
Forest plot of composite vascular events at 90 days in acute ischemic stroke patients receiving minocycline compared with placebo.

#### Stroke Recurrence at 90 Days

Stroke recurrence at 90 days was examined in three studies enrolling 2,014 patients. We found no statistically significant difference between minocycline and control (OR 0.99, 95% CI 0.65 to 1.45; Z=0.03, P=0.98; moderate certainty). Heterogeneity was absent (I^2^=0%, Chi^2^=1.68, df=3, P=0.64), The forest plot for stroke recurrence is displayed in Figure 8.

**Figure 8.**
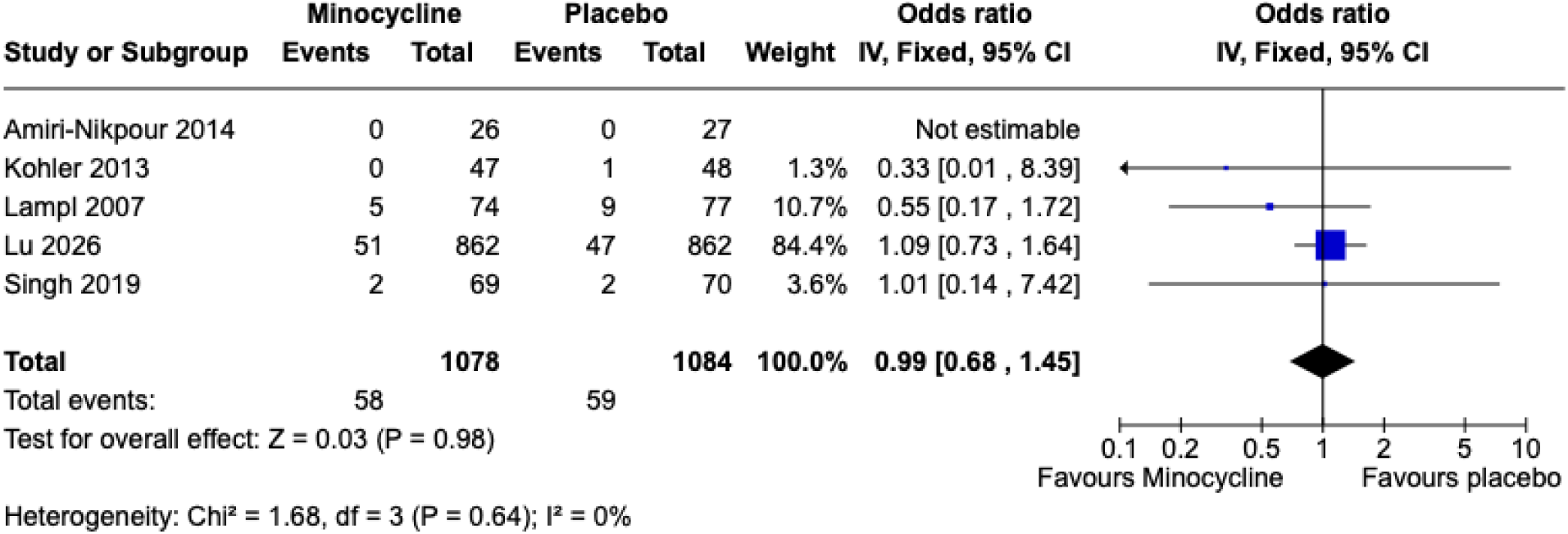
Forest plot of stroke recurrence at 90 days in patients treated with minocycline compared with control following ischemic stroke.

### SUBGROUP ANALYSES

#### Stroke type

Pre-specified subgroup analyses examined whether the effect of minocycline differed between AIS, ICH, and mixed AIS/ICH populations for key neurological and functional outcomes. Detailed estimates for NIHSS, mean mRS, mRS 0–2, and mRS 0–1 at 90 days by stroke type are summarized in Supplementary Table 2, with corresponding forest plots provided in Figure 9.

**Figure 9.**
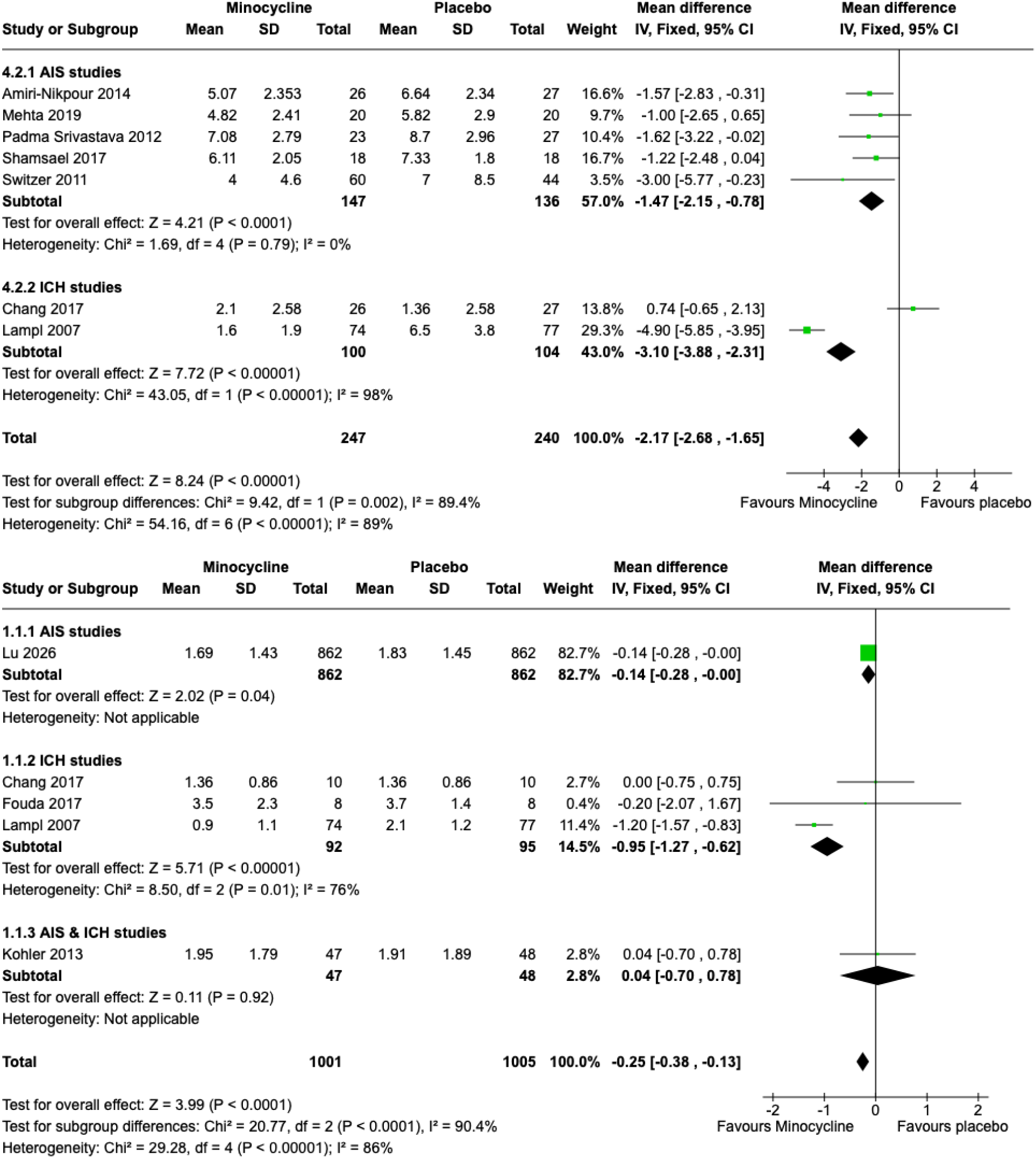
Forest plots showing subgroup analyses by stroke type. Upper panel: mean difference in NIHSS score at 90 days for minocycline versus placebo in AIS and ICH studies. Lower panel: mean difference in modified Rankin Scale score at 90 days for minocycline versus placebo across AIS, ICH, and mixed AIS/ICH studies. AIS: Acute ischemic stroke, ICH: Intracerebral Hemorrhage, NIHSS: National Institutes of Healt Stroke Scale

In this analysis, AIS-only cohorts consistently showed benefit across outcomes, whereas ICH and mixed AIS/ICH subgroups were based on smaller, more heterogeneous datasets and yielded more variable effect sizes. Tests for interaction indicated that stroke type significantly modified the effect on NIHSS and mean mRS, but not on dichotomized functional outcomes (mRS 0–2 or 0–1). These subgroup findings should, therefore, be interpreted as exploratory and are presented primarily to inform future hypothesis-driven trials in AIS and ICH, rather than to support stroke-type-specific treatment recommendations at this stage.

#### Route of administration

We also evaluated whether the effect of minocycline differed between oral and intravenous administration. Pooled estimates for NIHSS, mean mRS, mRS 0–2, and mRS 0–1 at 90 days by route are summarized in Supplementary Table 3, with full forest plots presented in Figures S5–S8.

Across outcomes, orally treated cohorts generally showed directionally consistent and statistically favorable effects, whereas intravenous-only studies were fewer, smaller, and yielded imprecise estimates around the null. However, formal tests for interaction by route were not statistically significant for the dichotomized functional outcomes, and the overall conclusions of the meta-analysis were unchanged when analyses were stratified by administration route. Accordingly, these findings should be viewed as hypothesis-generating and primarily informative for the design of future trials comparing specific oral versus intravenous regimens rather than as definitive evidence in favor of one route over the other.

## DISCUSSION

This systematic review and meta-analysis, comprising 11 studies and 2,428 patients, provides the most comprehensive quantitative synthesis to date of minocycline as an adjunctive neuroprotective strategy in acute stroke. In both the primary and secondary outcomes, minocycline was consistently linked to better neurological and functional recovery at 90 days, including a significant drop in NIHSS score, a lower mean mRS, and higher odds of excellent functional independence defined as mRS 0–1. These findings extend and substantially reinforce the conclusions of the 2018 meta-analysis by Malhotra et al. (11), which was restricted to 426 patients from randomized trials alone and could not draw definitive conclusions due to limited statistical power. These results are also directly corroborated by the recently published EMPHASIS trial, the largest randomized controlled trial of minocycline in AIS to date, which demonstrated that minocycline initiated within 72 hours significantly improved the probability of an excellent functional outcome (mRS 0–1) at 90 days. (12)

### Biological Plausibility and Mechanistic Framework

The clinical benefit observed across multiple outcomes in the present analysis is biologically coherent with minocycline’s well-characterized pleiotropic neuroprotective mechanisms. After the initial ischemic insult, secondary neurological injury is propagated through a cascade involving microglial activation, neuroinflammation, programmed cell death, BBB disruption, and MMP-mediated extracellular matrix degradation.(3,4) Minocycline attenuates each of these pathways: it suppresses microglial M1 activation and promotes polarization toward the neuroprotective M2 phenotype through modulation of the STAT1/STAT6 signaling axis,(24,25) reduces neuronal apoptosis via inhibition of cytochrome c release and caspase-3 activation, and inhibits MMP-9 activity, thereby preserving BBB integrity and reducing the risk of hemorrhagic transformation. In the MINOS trial, IV minocycline produced a dose-dependent reduction in plasma MMP-9 levels in both thrombolysis-treated and non-thrombolysis-treated patients, providing direct in-human mechanistic evidence linking minocycline to one of its primary therapeutic targets.(37) The neuroprotective effect on ICH models, where minocycline reduces extracellular matrix metalloproteinase inducer (EMMPRIN) and MMP-9 expression, decreases brain edema, and limits perihematomal neuroinflammation, may also explain the larger treatment effects observed in ICH subgroups, despite the limited sample sizes in those cohorts.(26)

### Primary Outcome: Neurological Recovery

The primary outcome of neurological recovery demonstrated a clinically meaningful improvement. A 2-point improvement in NIHSS has been validated as a threshold associated with a measurable reduction in functional dependency in multiple stroke outcome studies, and the lower bound of the confidence interval (−1.65) remains above this threshold even under the most conservative estimate.(12) Importantly, this effect proved robust to prespecified sensitivity analyses and to stratification by stroke type, with consistent benefit in AIS and a plausible but less certain signal in ICH. The consistent direction of effect across five independent AIS trials in the subgroup analysis (10,29,30,35,36) further supports the robustness of this finding. Although between-study heterogeneity for NIHSS was substantial, much of this could be attributed to one early high-impact trial and to the small, methodologically heterogeneous ICH cohorts (28,33), suggesting that the observed neurological benefit reflects a genuine treatment effect rather than an artifact of a single outlier study.

### Functional Outcomes and the EMPHASIS Trial’s Contribution

The consistent benefit observed across multiple functional outcome metrics: mean mRS, mRS 0–2, mRS 0–1, and composite vascular events, substantially strengthens the evidence for minocycline’s efficacy. The low heterogeneity for mRS 0–1 (I^2^=0%) and composite vascular events (I^2^=0%) is particularly notable, as it indicates that the functional benefit is reliably consistent across the contributing studies despite their methodological diversity. The magnitude of these effects was small to moderate at the individual-patient level but is similar to that reported for other neuroprotective candidates (38). The mRS 0–2 outcome reached only borderline significance (OR 1.21, 95% CI 1.00 to 1.47; P=0.05), which may reflect both the more stringent definition of ‘excellent outcome’ (complete or near-complete recovery) and the heterogeneity introduced by the Padma Srivastava (35) and Singh (31) trials with opposing estimates. In all three functional outcome analyses, the EMPHASIS trial (12) contributed >80% of total pooled weight, meaning that the pooled estimates are heavily anchored by this high-quality multicenter trial. Leave-one-out exclusion of EMPHASIS (12) still showed a significant reduction in disability, suggesting that smaller trials are directionally concordant, even though the overall estimate is highly influenced by this large phase 3 trial. Subgroup-based sensitivity analyses by stroke type and route of administration did not alter the conclusions: benefit remained evident in AIS and in orally treated cohorts, whereas ICH-specific and IV-only estimates were imprecise and should be regarded as exploratory. While this introduces sample domination, it is arguably a strength rather than a limitation: the presence of one large methodologically rigorous trial providing directionally consistent results with smaller independent studies is precisely the evidentiary structure that underpins confidence in a treatment effect.(12)

### Null Findings: Stroke Recurrence

By contrast, minocycline did not reduce 90-day stroke recurrence, and the composite vascular endpoint, although statistically favorable, should be interpreted cautiously. From a mechanistic perspective, this dissociation is not surprising. The biological rationale for minocycline in stroke is centered on modulation of neuroinflammation, apoptosis, and BBB injury in the ischemic penumbra or perihematomal region, rather than on modification of the underlying atherothrombotic or cardioembolic substrate that determines recurrent event risk (5,6). The limited number of studies contributing to vascular endpoints and the wide confidence intervals around individual estimates mean that the current data are insufficient to claim either a robust vascular protective effect or any signal of harm (28,31). Instead, they are most consistent with minocycline acting as a neuroprotective adjunct that improves recovery from the index event while leaving recurrent vascular risk largely unchanged under contemporary secondary prevention strategies (11).

### Subgroup Heterogeneity by Stroke Type

The significant interaction by stroke type observed for NIHSS (P=0.002) and mean mRS (P<0.0001) deserves further consideration. The numerically larger effect in ICH studies is mechanistically plausible (Figure 9): perihematomal neuroinflammation, microglial activation, BBB disruption, and edema formation represent quantitatively dominant contributors to secondary injury in ICH compared to AIS, and these are precisely the pathways most potently inhibited by minocycline.(34,37) However, the ICH evidence base in this meta-analysis is severely limited: only two to three studies contributed to ICH subgroups, with extreme within-subgroup heterogeneity and small sample sizes ranging from 16 to 151 patients. The absence of a significant subgroup difference for the dichotomized mRS 0–2 and mRS 0-1 outcome (P=0.66) further suggests that the continuous outcome (NIHSS and mRS mean difference) interaction may be partially driven by scale effects and measurement heterogeneity rather than a truly differential biological response.

### Dose, Route, and Treatment Timing Considerations

Substantial variability in minocycline dosing, ranging from 200 mg/day orally to 10 mg/kg IV, and treatment initiation windows (6 to 72 hours) across included studies limit definitive conclusions about optimal regimen design from this meta-analysis alone. Dose-escalation data from the MINOS trial showed that intravenous minocycline is safe and well-tolerated up to 10 mg/kg, with a half-life of about 24 hours. (10) IV administration also achieves higher and more predictable peak plasma concentrations relevant to MMP-9 inhibition than oral dosing. (35) Conversely, the large EMPHASIS trial demonstrated clinically meaningful benefit with an oral loading-dose regimen, which offers substantial logistical and cost advantages in real-world implementation, particularly in low- and middle-income settings. (12) The observation that benefit was preserved even with treatment initiation up to 72 hours (12) is clinically important, as it suggests that minocycline’s anti-inflammatory and anti-apoptotic mechanisms remain relevant well beyond the reperfusion window, a feature that distinguishes it from time-sensitive reperfusion therapies and broadens its potential reach to a substantially larger proportion of the global stroke population. (27)

### Safety Profile

Minocycline demonstrated a favorable safety profile across the included studies. Rates of symptomatic intracranial hemorrhage, serious adverse events, vascular death, and all-cause mortality at 90 days were low and comparable between minocycline and control arms in all trials that reported safety data, including EMPHASIS (vascular death 1.2% vs. 1.9%; all-cause death 1.6% vs. 2.3%). (12) The MINOS dose-escalation trial specifically confirmed tolerability at doses up to 10 mg/kg IV in combination with IV tPA, with no excess of symptomatic hemorrhagic transformation. (10) These safety data are reassuring and consistent with minocycline’s established tolerability profile in non-stroke indications. The theoretical concern regarding antibiotic-related adverse effects, including gastrointestinal intolerance and cutaneous reactions, was not systematically evaluated across all studies.

### Strengths

This meta-analysis has several notable strengths. First, with 2,428 patients across 11 studies, it is the largest quantitative synthesis of minocycline in acute stroke to date, providing substantially greater statistical precision than any individual study or prior meta-analysis. (11) Secondly, the prospective PROSPERO registration and adherence to PRISMA 2020 guidelines minimize post-hoc bias in outcome selection and analysis. Similarly, pre-specified subgroup analyses by stroke type and leave-one-out sensitivity analyses allowed for a systematic evaluation of heterogeneity sources and an assessment of result robustness. Also, the inclusion of the EMPHASIS trial, a landmark multicenter trial with 1,724 patients that was unavailable to prior syntheses, fundamentally changes the weight of available evidence and provides a high-quality anchor for the pooled estimates.(12) Finally, the inclusion of both AIS and ICH studies, while contributing to heterogeneity, allows broader evaluation of minocycline’s potential across stroke subtypes and generates hypothesis-generating evidence for stroke-type-specific trials.

### Limitations

Several limitations must be acknowledged; firstly, substantial statistical heterogeneity was present for continuous outcomes (I^2^=86–89%), reflecting the clinical diversity of included populations in terms of stroke severity, stroke subtype, treatment regimen, background reperfusion therapy, and geographic setting. Although sensitivity analyses confirmed the robustness of the primary findings, the pooled estimates should be interpreted as average effects across heterogeneous settings rather than precise predictions for any specific patient population. Moreover, the EMPHASIS trial contributes >70% of the total sample and >80% of the weight in functional outcome analyses, making the pooled estimates heavily dependent on a single trial. In fact, nine of the eleven included studies were conducted in Asia and the Middle East, and the generalizability of findings to Western populations with different genetic backgrounds, comorbidity profiles, and stroke management practices may be limited. Besides, outcome definitions and follow-up intervals varied across studies; for example, MINOS reported only 7-day outcomes, and the measurement of mean NIHSS at 90 days as a continuous variable (rather than absolute change from baseline) was not uniformly reported, requiring standard statistical imputation in some cases. Lastly, publication bias cannot be fully excluded given the small-study effects observed in several subgroup analyses and systematic evaluation of funnel plot asymmetry were not possible for all outcomes due to the limited number of contributing studies.

### Future Directions

Several key evidence gaps identified by this meta-analysis should inform the design of future trials. Firstly, adequately powered trials dedicated to ICH are urgently needed: the present synthesis provides only a hypothesis-generating signal for minocycline in ICH, and the extreme heterogeneity between Lampl (28) and MITCH (33) illustrates that ICH-specific trials require rigorous blinding, larger sample sizes, and standardized outcome definitions. Similarly, optimal dosing, particularly the question of whether IV or oral loading-dose strategies produce superior outcomes compared to the standard 200 mg/day oral regimen, remains unanswered and would benefit from adaptive dose-finding designs. Third, a confirmatory multicenter trial evaluating minocycline in combination with IV thrombolysis is warranted, building on MINOS safety data demonstrating tolerability with tPA and preclinical evidence suggesting that minocycline may extend the therapeutic window for thrombolytic therapy by reducing hemorrhagic transformation risk. (10,37) Such a trial is currently ongoing (ClinicalTrials.gov NCT07526987). (39) Fourth, biomarker-enriched trial designs incorporating imaging-based penumbral selection or circulating neuroinflammatory markers could reduce pathophysiological heterogeneity and improve the proportion of ‘informative’ patients in future neuroprotective trials, addressing a recognized cause of prior neuroprotection failures.(40)

## CONCLUSIONS

This meta-analysis of 11 studies and 2,428 patients demonstrates that minocycline is associated with significant improvements in 90-day neurological and functional outcomes in acute stroke, with a consistent direction of benefit across NIHSS, mean mRS, mRS 0–2, and composite vascular events. The low cost, oral bioavailability, broad therapeutic window, established safety record, and global availability of minocycline distinguish it from virtually all other candidate neuroprotective agents and position it as a potentially scalable adjunctive therapy of particular relevance in resource-limited settings. The totality of the available evidence — anchored by the landmark EMPHASIS trial and supported by consistent directional findings across independent smaller trials — supports the prioritization of minocycline for confirmatory trials and justifies its evaluation as a candidate for guideline consideration in AIS management.

## Data Availability

The datasets generated and analyzed during the current study are available from the corresponding author upon reasonable request.

## Abbreviations

AIS: Acute Ischemic Stroke
BBB: Blood-Brain Barrier
BI: Barthel Index
CI: Confidence Interval
CSSRS: Columbia Suicide Severity Rating Scale
DALY: Disability-Adjusted Life Year
df: Degrees of Freedom
EMMPRIN: Extracellular Matrix Metalloproteinase Inducer
EQ-5D: EuroQol 5-Dimension
ERK1/2: Extracellular signal-Regulated Kinase 1/2
GCS: Glasgow Coma Scale
GRADE: Grading of Recommendations Assessment, Development, and Evaluation
I^2^: I-squared
ICH: Intracerebral Hemorrhage
IQR: Interquartile Range
IV: Intravenous
M1/M2: Microglial polarization phenotypes
mBI: modified Barthel Index
MD: Mean Difference
MI: Myocardial Infarction
MMP: Matrix Metalloproteinase
MMP-9: Matrix Metalloproteinase-9
mRS: modified Rankin Scale
NIHSS: National Institutes of Health Stroke Scale
NR: Not Reported
OR: Odds Ratio
PI3K: Phosphoinositide 3-Kinase
PRISMA: Preferred Reporting Items for Systematic Reviews and Meta-Analyses
PROBE: Prospective, Randomized, Open-label, Blinded Endpoint
PROSPERO: International Prospective Register of Systematic Reviews
RCT: Randomized Controlled Trial
RoB 2: Cochrane Risk of Bias tool version 2
ROBINS-I: Risk Of Bias In Non-randomised Studies of Interventions
rt-PA/tPA: recombinant tissue Plasminogen Activator
SD: Standard Deviation
SRA: Systematic Review Accelerator
STAT1/STAT6: Signal Transducer and Activator of Transcription 1/6
TIA: Transient Ischemic Attack

## Author Contributions

Conceptualization, ML and ND; methodology, ML, AH and YS; software, ML, ND, MG, MR and YS; validation,

ML, AH and NS; formal analysis, ML, AH, YS; investigation, AH, ND, YS and ML; resources, ND, ML; data curation, ML, ND, YS; writing—original draft preparation, ML, ND, YS; writing—review and editing,

ML, NS and SD; visualization, SD, and ML; supervision, ML, SD and NS; project administration, ML.

All authors have read and agreed to the published version of the manuscript.

## Ethics Approval

Not applicable. This study is a systematic review and meta-analysis of published data and does not involve human or animal subjects.

## Consent to Participate

Not applicable.

## Data Availability

All data generated or analyzed during this study are included in this published article and its supplementary information files.

